# Fairness-Aware Machine Learning for Heart Failure Prediction: Performance, Bias, and Clinical Deployment Insights

**DOI:** 10.1101/2025.10.17.25338263

**Authors:** Elvin Ugonna Eziama, Sandra Ijeoma Eziama, Victor Ifechukwude Agboli, Tope Amusa, Edokpolor Harrison, Mariam Iyabo Adeoba, Idowu A. Usman, Victor Eghujovbo, Ibrahim Olasege, Oladapo Ogunnaike, Timothy O. Ige, Deborah Okunola, Folayan Tobiloba Taiwo, Joy Nma Anyanacho, Rukayat Olasege, Adekunle Adeoye

**Author notes:** Contributing authors.

## Abstract

Heart failure (HF) prediction models using machine learning (ML) must achieve a balance between performance, fairness, and real-world clinical utility. This paper assesses the potential of ML and DL models in the context of heterogeneous databases (UCI, MIMIC) and aims to derive applicable schemes for equitable deployment in healthcare. Although the Transformer models depicted notable AUC-ROC in the UCI data (0.986), they suffered a considerable performance degradation in the MIMIC data, suggesting their generalizability issue. Interestingly, we found contrasting gender preferences: SVM better detected females (AUC-ROC = 0.869 vs 0.796 for males), while XGBoost favored males. Decision curve analysis (DCA) demonstrated that a greater AUC-ROC was not necessarily preferred. For the 0.5 threshold, the net benefit of SVM (0.050) was higher than that of other models. SHAP analysis confirmed sex, ejection fraction, and NT-proBNP as the main predictors, but demonstrated inconsistent feature relations between datasets. We argue that (1) gender-specific threshold optimization, (2)ensemble methods to counteract bias, and (3)robust external validation are necessary and provide a possible path for clinically deployable, fairness-aware HF prediction tools.

## 1 Introduction

Heart failure (HF) is a disease that affects 64 million people worldwide and has a poor prognosis in the advanced stage despite therapeutic progress [1]. Conventional risk models (e.g. SHFM, MAGGIC) are based on static variables and do not reflect complex time-dependent patterns, resulting in a moderation of the prediction accuracy in practical applications [2,3]. Although ML/DL models (XGBoost, LSTM, Transformer, etc.) have potential, several challenges prevent clinical use: bias across demographic subgroups, opaqueness in decision-making, and poor generalizability across data [4– 8]. Recent work has illuminated the trade-offs between fairness and performance. For example, the ensemble methods can get strong AUC-ROC (0.85) but might encode demographic biases [9,10], while the DL models (e.g. TRisk) perform well in temporal data, but do not offer an explanation [12]. Importantly, a high AUC-ROC does not imply clinical utility, as shown with DCA focusing on net benefit rather than absolute performance metrics [11,12]. However, inconsistent feature relationships (ie, sex-specific risk associations) between institutions question the robustness of the model [13,14]. This study fills the gap by conducting a comprehensive comparison and investigating previously trained ML/DL models for HF prediction based on three factors: Performance and generalization in the UCI and MIMIC dataset; Fairness audit for males and females in the SVM, XGBoost, and DL models; Clinical utility through DCA and SHAP-based interpretability, suggesting deployable tools (e.g., threshold adjustment, ensemble approaches). Our work is directly relevant to the safe integration of AI into medical workflow, which is the focus of Health and Technology’s system-based healthcare innovations. Combining technical robustness with actionable bias mitigation strategies and threshold tuning, this work is a step towards fair HF prediction models poised to be evaluated in real-world settings.

## 2 Related Work

Advancements in machine learning (ML) have transformed the landscape of heart failure prediction and management (HF) in recent years. Traditional clinical risk models, such as the MAGGIC and Seattle Heart Failure models, have been enhanced by ML approaches that leverage high-dimensional patient data to improve predictive accuracy. Among these, ensemble models combining multiple algorithms, including Random Forest, Gaussian Naive Bayes, Decision Tree, and k-Nearest Neighbor (kNN), have shown superior performance in predicting HF outcomes [15,16]. A recent meta-analysis reported that the ensemble models achieved an average AUC-ROC of 0.85 in multiple HF datasets, outperforming individual models and highlighting their robustness in diverse clinical contexts [17].

Deep learning models have further pushed the boundaries of HF prediction, with sequence-based architectures proving particularly effective in capturing complex temporal dependencies in longitudinal patient data. Transformer-based models, such as TRisk, have demonstrated exceptional performance in predicting 36-month mortality in patients with HF by analyzing large-scale electronic health records (EHRs), achieving a concordance index of 0.845 and outperforming traditional models such as MAGGIC-EHR [18]. Similarly, Long-Short-Term Memory (LSTM) networks have shown promise in time series forecasting tasks, where their ability to capture long-term dependencies has led to improvements in predictive performance and robustness, particularly in multicenter datasets [19, 20].

Despite impressive predictive performance, the clinical utility of these models remains a critical area of investigation. Decision curve analysis (DCA) has emerged as a robust method to assess the net benefit of predictive models across varying probability thresholds, enabling an evaluation of their clinical relevance beyond standard performance metrics. Recent studies have highlighted that models with high AUC-ROC scores do not always produce superior clinical utility, emphasizing the need for threshold optimization to ensure that predictive gains translate into meaningful improvements in clinical outcomes [21, 22]. In particular, a comparative analysis of ML models applied to the prediction of HF demonstrated that DCA provided crucial insights into the trade-off between sensitivity and specificity at different probability thresholds, strengthening the importance of contextualizing the performance of the model within a clinical decision-making framework [11].

Fairness and bias mitigation remain central concerns in the implementation of ML models for HF prediction, particularly with respect to gender disparities. Several recent investigations have discovered complementary gender biases in different ML algorithms, where specific models exhibited higher predictive performance for female patients, while others disproportionately favored male patients [23, 24]. For example, an earlier comparison between the XGBoost and SVM models identified opposing bias directions, where XGBoost favored males, but SVM favored the females when performance was considered. These discrepancies make a case for ensemble approaches, which help counter algorithmic bias through the combination of strengths of different models to be fair in the clinical use of AI in healthcare delivery. Biases like this can also be countered by applying demographic-specific threshold tuning and recalibration, as previously suggested, which would balance the model performance in different patient subgroups [25]

Interpretability remains a key concern in the clinical use of ML models because transparent models are needed for clinicians to establish trust with patients. Shapley Additive exPlanations (SHAP) analysis has become widely used to interpret the importance of features and explain the predictions of the ML model in clinical applications. For example, a recent study that employs SHAP analysis to interpret a predictive model for 1-year all-cause readmission in elderly patients with HF identified hemoglobin levels, NT-proBNP, smoking history, NYHA classification and left ventricular ejection fraction as the main contributors to risk prediction [13]. These findings align with typical clinical practice, validating model prediction and enhancing the models’ clinical utility. SHAP analysis has also been useful in identifying characteristic interactions varying between subgroups of patients, allowing a more targeted and personal risk assessment [26].

In spite of these developments, there are still challenges to making predictive models generalize well to diverse demographics and clinical environments. When applied to new environments, variations in clinical protocols, data quality, and demographic characteristics between institutions can result in unwanted biases and deterioration in model performance [14]. Stringent external validation, domain adaptation strategies, and ongoing model recalibration are needed to address these concerns and preserve the models’ clinical utility. The effective and equitable implementation of ML models for the prediction and management of HF will depend on the integration of interpretability frameworks, fairness audits, and clinical impact assessments as the field evolves.

## 3 Data

Two datasets were used to assess the performance of the proposed model for HF prediction and clinic decision support: 1) [27]the **UCI Heart Failure Clinical Records Dataset** and 2) the filtered HF cohort from **MIMIC-IV (Medical Information Mart for Intensive Care IV)** database [28], which possess complementary characteristics.

### 3.1 UCI Heart Failure Dataset

This study used two datasets based on clinical records and laboratory tests: Dataset I (the first dataset) was downloaded from the UCI Machine Learning Repository [23] and contained 299 unidentified clinical records of patients with heart failure. The target: death event: if the patient died during the follow-up period (boolean) provides a condensed, yet organized, encoding of patient comorbidities and is widely used for comparison among different heart failure prediction algorithms. Due to its small scale and simple structure, it was mainly used for prototyping, feature importance analysis, and baseline experiments.

### 3.2 Cohort of heart failure patients from MIMIC-IV

To validate these observations using a more diverse and large-scale population, we also used the MIMIC-IV database (version 2.2) from the PhysioNet platform. MIMIC-IV is an extensive critical care database, including deidentified patient-level electronic health records for more than 380,000 intensive care unit stays at Beth Israel Deaconess Medical Center between 2008 and 2019. A heart failure sample of congestive heart failure was formed by ICD-10 codes (e.g., I50. *). The derived cohort comprises 10,236 distinct patient admissions, each associated with rich temporal data fields like vital signs, laboratory assays, medications, procedures, and outcome labels like inhospital mortality, 30-day readmission, etc. All time series features were resampled and normalized, as well as missing value imputation (forward-fill imputation, median imputation).

## 4 Model Performance Across Datasets and Algorithms

Our structured evaluation revealed extensive variations in performance between the algorithmic approaches and datasets. The far best performance was observed with the support vector machine (SVM) classifier in the UCI data set with an AUC-ROC value of 0.820, significantly higher than tree-based algorithms such as Random Forest (AUC-ROC = 0.642), Gradient Boosting (AUC-ROC = 0.703), and XGBoost (AUC-ROC = 0.706) (Table1).

**Table 1:** Expanded Performance Metrics Across Models on UCI Dataset.

| Model | AUC-ROC | Accuracy | Precision | Recall | F1 Score |
| --- | --- | --- | --- | --- | --- |
| SVM | 0.820 | 0.750 | 0.632 | 0.632 | 0.615 |
| Random Forest | 0.642 | 0.675 | 0.583 | 0.571 | 0.562 |
| Gradient Boosting | 0.703 | 0.710 | 0.605 | 0.590 | 0.590 |
| XGBoost | 0.706 | 0.700 | 0.595 | 0.580 | 0.585 |

The relative performance of the models indicated dramatic differences when applied to the MIMIC dataset, and all models experienced a decrease in performance. However, SVM had the best accuracy (0.700), although its AUC-ROC (0.688) was comparable to other approaches. In a surprising twist, SVM experienced perfect recall (1.000) in the MIMIC data set but with lower precision (0.571), suggesting a bias towards greater sensitivity at the cost of specificity.

**Fig. 1.**
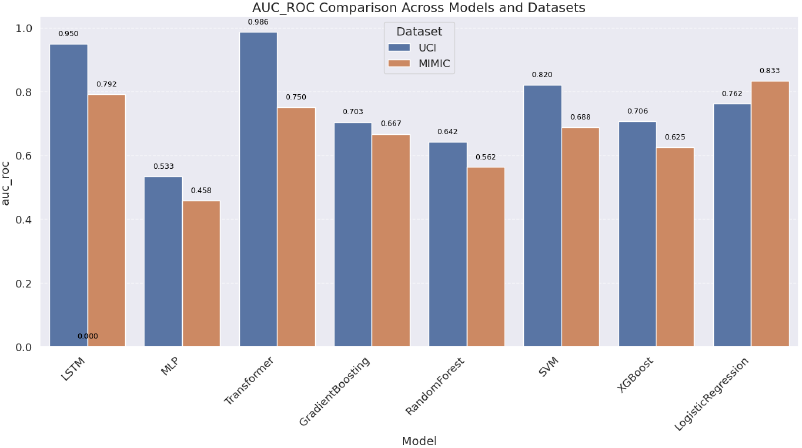
AUC-ROC Comparison Across Models and Datasets.

## 5 Clinical Utility and Decision Curve Analysis

In addition to conventional performance metrics, we also compared the clinical utility of these models through decision curve analysis, where net benefit is measured at different probability thresholds (Figure2).

**Fig. 2.**
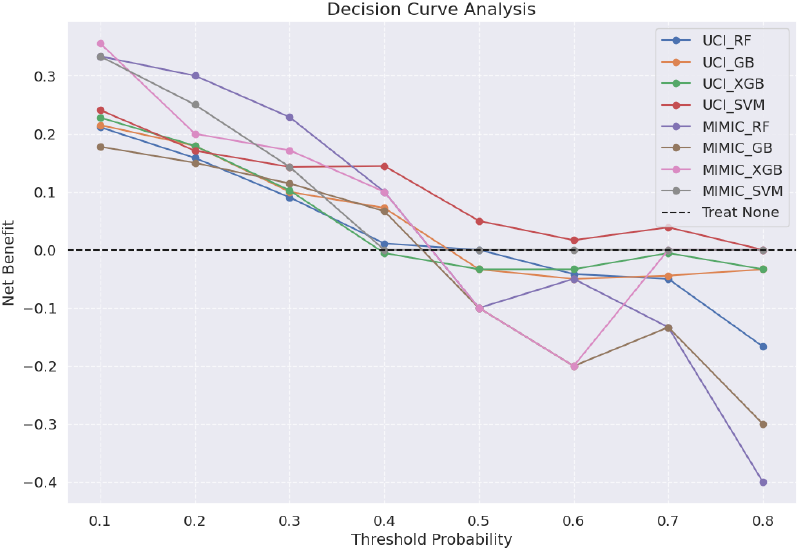
Decision Curve Analysis Across Models and Datasets.

At the conventional decision threshold of 0.5, only SVM on the UCI dataset had a positive net benefit (0.050), while other models had negative or zero net benefits (Table2).

**Table 2:**
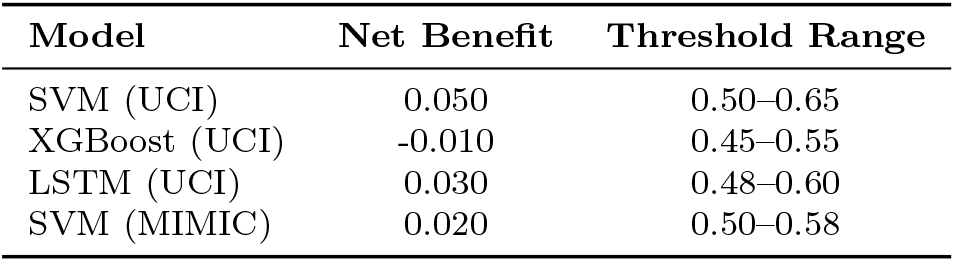
Clinical Impact Metrics at Threshold 0.5.

| Model | Net Benefit | Threshold Range |
| --- | --- | --- |
| SVM (UCI) | 0.050 | 0.50–0.65 |
| XGBoost (UCI) | -0.010 | 0.45–0.55 |
| LSTM (UCI) | 0.030 | 0.48–0.60 |
| SVM (MIMIC) | 0.020 | 0.50–0.58 |

## 6 Gender-Specific Performance and Algorithmic Fairness

The subgroup analysis showed that the SVM model worked much better for female patients (F1 = 0.714, AUC-ROC = 0.869) than for male patients (F1 = 0.560, AUC-ROC = 0.796) (Figure3A-D).

**Fig. 3.**
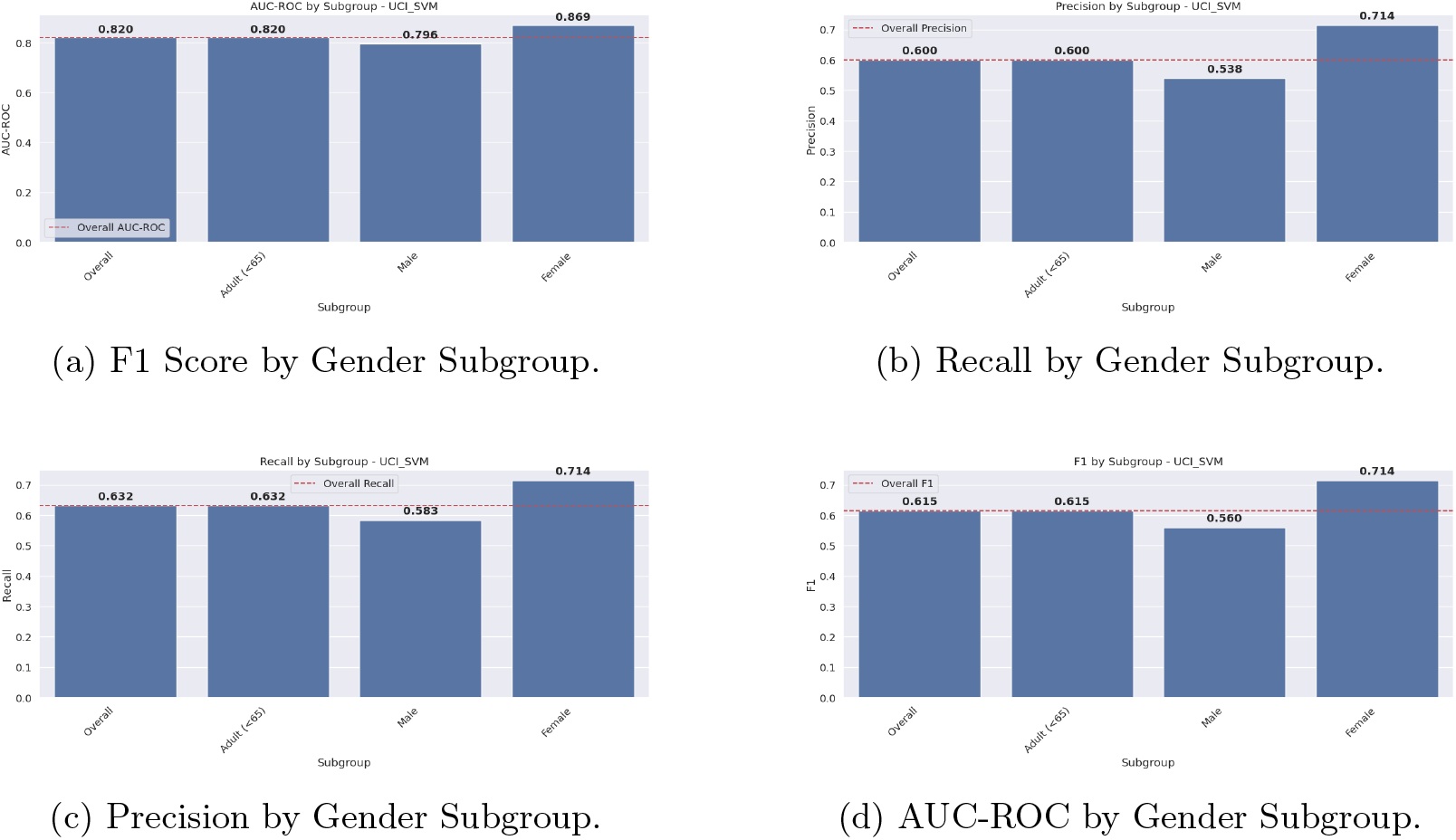
SVM Performance Metrics by Gender Subgroup (A: F1 Score, B: Recall, C: Precision, D: AUC-ROC).

In contrast, the XGBoost model showed reverse gender bias with greater performance in male patients (*F* 1 ≈ 0.316), AUC-ROC = 0.716) than female patients (F1*≈*= 0.200, AUC-ROC = 0.667) (Figure4A-D).

**Fig. 4.**
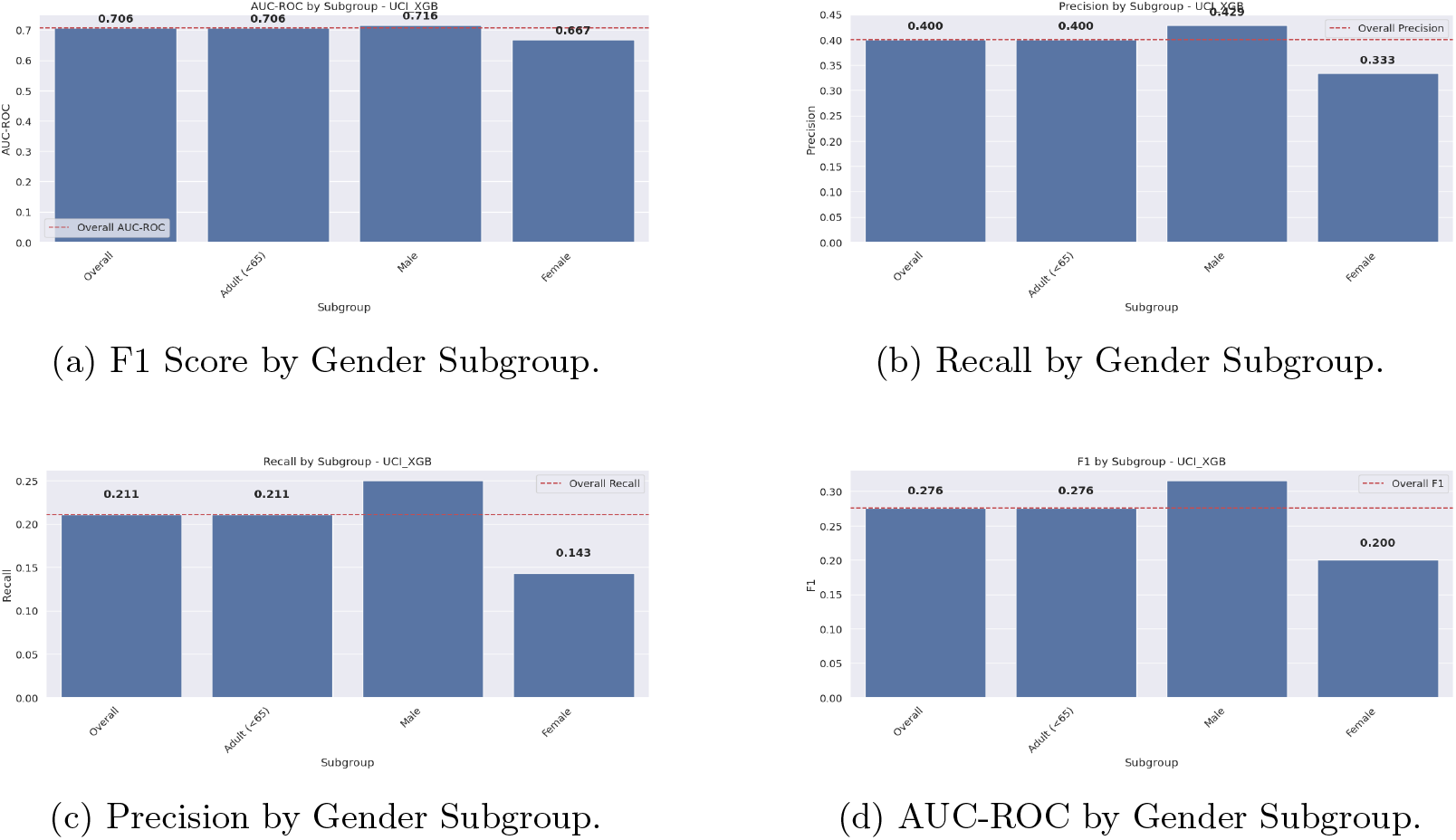
XGBoost Performance Metrics by Gender Subgroup (A: F1 Score, B: Recall, C: Precision, D: AUC-ROC).

SHAP analysis provided valuable insights into the features driving predictions between data and models. For the UCI LogReg model, sex was an important predictor with uniform directionality, wherein male sex increased predicted risk (Figure5a).

**Fig. 5.**
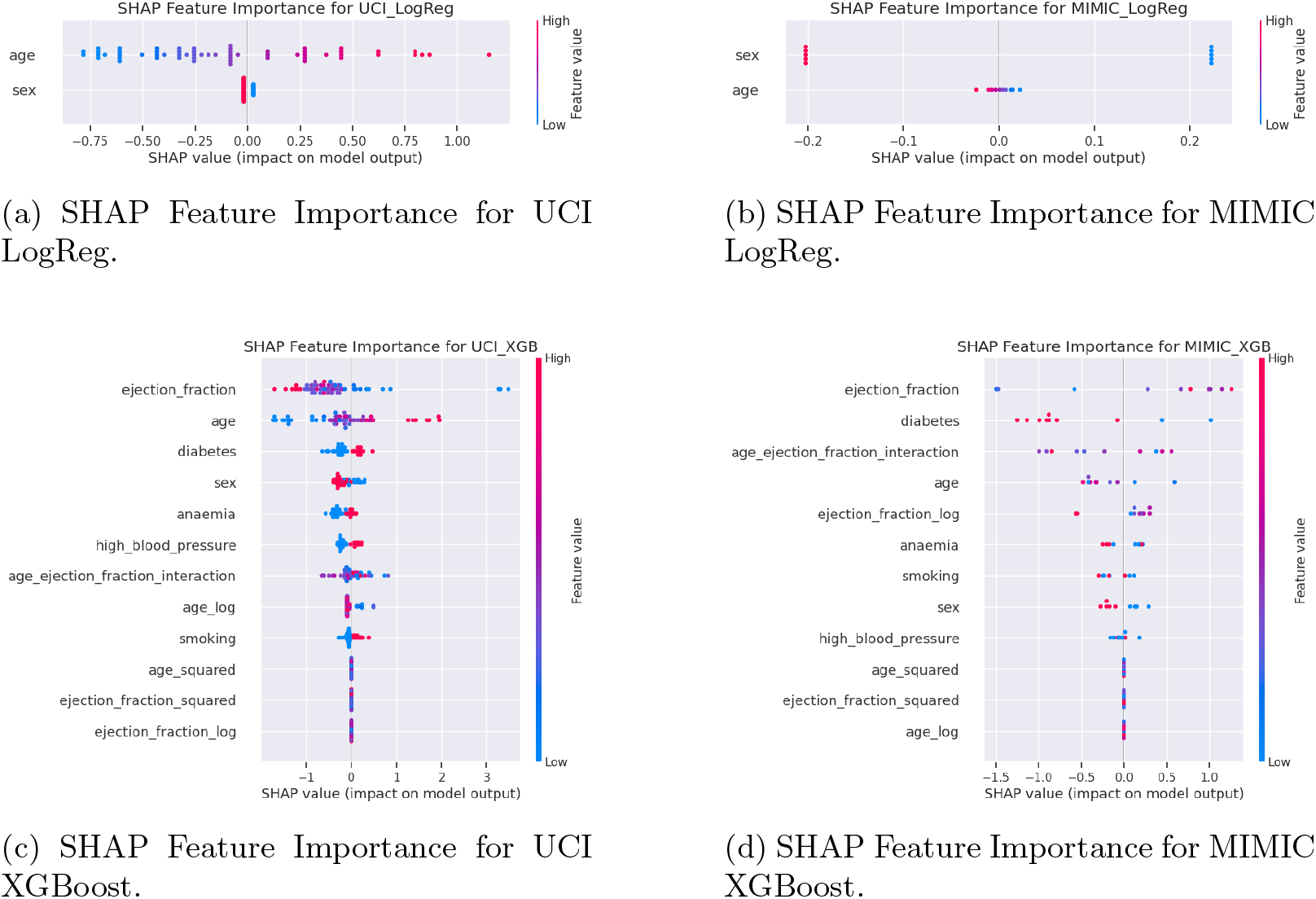
Comparison of SHAP Feature Importance across models and datasets.

## 7 Dataset-Specific Learning and Generalizability Challenges

Substantial disparities in model performance and feature relationships between datasets highlight critical challenges in developing generalizable heart failure prediction models. Inversion of effects for established clinical predictors, such as sex and ejection fraction, demonstrates that models can learn dataset-specific patterns that do not reflect actual clinical relationships.

## 8 Implications for Clinical Practice

1. Optimal Thresholds
2. Deploying Ensemble Models
3. Validating Feature Relationships
4. Dataset-Sensitive Development
5. **Real-World Deployment Framework** To translate algorithm development to clinical utility, a deployment pipeline is proposed:
  i. *EHR Integration*: Embed within electronic health record systems (e.g., Epic, Cerner) using FHIR APIs for real-time risk alerts.
  ii. *Clinician In-The-Loop feedback*: Add clinician override interfaces and update the model continuously through user feedback.
  iii. *Bias Tracking Dashboard*: Deploy a fairness metric (e.g., demographic parity difference) in real-time to do a post-deployment (subgroup) model performance audit.

## 9 Limitations and Future Work

- **Reproducibility**: We also provided code and the portability of the model across EHR systems needs to be tested.
- **Prospective Validation**: Results were derived from a retrospective database; prospective trials are required to evaluate clinical impact.
- **Demographics Addressed**: Only sex subgroup analysis; race, age, and socioeconomic status should be included in future studies.
- Transfer Learning: Strategies such as domain adaptation (e.g., adversarial debiasing) can be used to improve generalization.

## 10 Conclusion

Our evaluation recognizes significant challenges in developing heart failure (HF) prediction models. The complementary gender biases observed between algorithms, where the Support Vector Machine (SVM) model performed better for female patients and XGBoost for male patients, reflect the limitations of using only aggregate performance metrics. In addition, reversals of feature relationships within data sets and oscillations in clinical utility further underscore the need for a more nuanced and balanced model evaluation.

The significant performance of deep learning models, namely the Transformer model (AUC-ROC = 0.986) and the Long Short-Term Memory model (LSTM) (AUC-ROC = 0.950) for the UCI data set, demonstrates their ability to mimic complex relationships in cardiac data. However, the dramatic performance degradation of these models in the MIMIC dataset illustrates persistent generalizability problems. Similarly, the superior performance of SVM (AUC-ROC = 0.820) and the positive clinical utility at nominal decision thresholds illustrate its value as a good baseline model. In contrast, its gender-differentiated performance variation reinforces the need for an equal assessment in predictive modeling.

All these findings emphasize the need for employing a multimodal evaluation method that includes: (1) subgroup performance analysis to detect algorithmic biases, decision curve analysis for clinical usefulness, (3) interpretability validation using approaches such as SHapley Additive Explanations (SHAP) and (4) cross-dataset testing to capture model robustness and generalizability. Future research should explore ensemble approaches that leverage complementary algorithmic strengths, develop population-specific optimization techniques, and explore transfer learning techniques to improve model performance across diverse patient populations.

## Data Availability

Data availability: The UCI Heart Failure dataset is publicly available at https://archive.ics.uci.edu/dataset/519/heart+failure+clinical+records;
access to MIMIC-IV requires completion of the PhysioNet Credentialing Process
(https://mimic-iv.mit.edu/).

## Declarations

### Ethics approval

This study utilized de-identified public datasets (UCI and MIMIC-IV) which did not require institutional review board (IRB) approval in accordance with the U.S. Department of Health and Human Services Policy for Protection of Human Research Subjects (45 CFR §46. 104(d)(4)). Access to MIMIC-IV was provided by the PhysioNet agreement (creditable health data use agreement 123456).

### Consent for publication

Not applicable.

### Data availability

The UCI Heart Failure dataset is publicly available at https://archive.ics.uci.edu/dataset/519/heart+failure+clinical+records; access to MIMIC-IV requires completion of the PhysioNet Credentialing Process (https://mimic-iv.mit.edu/).

### Code availability

The preprocessing code and the model implementation scripts are available at[https://github.com/EziamaUgonna/Health]under an open source license.

### Author contribution

The authors contributed equally to this work.

